# Chest CT versus RT-PCR for the Detection of COVID-19: Systematic Review and Meta-analysis of Comparative Studies

**DOI:** 10.1101/2020.06.22.20136846

**Authors:** Mohammad Karam, Sulaiman Althuwaikh, Mohammad Alazemi, Ahmad Abul, Amrit Hayre, Abdulmalik Alsaif, Gavin Barlow

**Author notes:** **Corresponding Author:** Mohammad Karam, Address: Al-Firdous, Block 4, Street 1, Avenue 6, Al-Farwaniyah, State of Kuwait.

## Abstract

**Objectives:** To compare the performance of chest computed tomography (CT) scan versus reverse transcription polymerase chain reaction (RT-PCR) as the reference standard in the initial diagnostic assessment of coronavirus disease 2019 (COVID-19) patients.

**Design:** A systematic review and meta-analysis were performed as per the Preferred Reporting Items for Systematic Reviews and Meta-analyses (PRISMA) guidelines. A search of electronic information was conducted using the following databases: MEDLINE, EMBASE, EMCARE, CINAHL and the Cochrane Central Register of Controlled Trials (CENTRAL).

**Setting:** Studies that compared the diagnostic performance within the same patient cohort of chest CT scan versus RT-PCR in COVID-19 suspected patients.

**Participants:** Thirteen non-randomised studies enrolling 4092 patients were identified.

**Main Outcome Measures:** Sensitivity, specificity and accuracy were primary outcome measures. Secondary outcomes included other test performance characteristics and discrepant findings between both investigations.

**Results:** Chest CT had a sensitivity, specificity and accuracy of 0.91 (0.82-0.98), 0.775 (0.25-1.00) and 0.87 (0.68-0.99), respectively, with RT-PCR as the reference. Importantly, early small, China-based studies tended to favour chest CT versus later larger, non-China studies.

**Conclusions:** A relatively high false positive rate can be expected with chest CT. It may still be useful, however, in patients with a suspicious clinical presentation of COVID-19 and a negative initial SARS-CoV-2 RT-PCR. In acute cardiorespiratory presentations, negative CT scan and RT-PCR tests is likely to be reassuring.

**Highlights:**

- The median accuracy of chest computed tomography (CT) for the diagnosis of coronavirus disease 2019 (COVID-19) is relatively high at 0.87 (range 0.68-0.99)
- Chest CT has relatively low specificity, even within the context of a pandemic, for the diagnosis of COVID-19 with an associated relatively high false positive rate
- Chest CT scan, however, appears to be able to detect most initially positive and most initially negative/subsequently positive RT-PCR diagnosed cases

## 1. Background

In December 2019, Coronavirus Disease 2019 (COVID-19), caused by Severe Acute Respiratory Syndrome Coronavirus 2 (SARS-COV-2) emerged from Wuhan city, China [1-2]. SARS-COV-2 is part of the Coronavirdae family that includes the common cold, Severe Acute Respiratory Syndrome (SARS) and Middle East Respiratory Syndrome (MERS). These viruses can cause Acute Respiratory Distress Syndrome (ARDS), bilateral pneumonia and pulmonary failure leading to mortality [3]. There is yet to be a highly effective pharmacological therapy identified for COVID-19, further highlighting the importance of early detection and isolation of COVID-19 patients to prevent the spread of the disease [4].

Reverse-transcription polymerase chain reaction (RT-PCR) tests are currently the gold standard diagnostic tool for COVID-19. RT-PCR assays can be performed on nasopharyngeal and/or oropharyngeal swabs, sputum, blood samples, body fluids, stool samples and bronchoalveolar lavage fluid [5]. RT-PCR is limited by its low sensitivity and associated relatively high number of false-negative results that can lead to the erroneous assumption that a patient who does actually have COVID-19 does not have it and is not infectious, leading to an undetected transmission risk in either the community or within an institutional setting [4].

CT has become a standard of care in the diagnosis and assessment of a variety of respiratory conditions and optimises the management process [6-7]. Although CT scans are not routinely used to diagnose ARDS, certain complications relating to mechanical ventilation, including pneumonia, pneumothorax and emphysema, are sometimes identified by CT but not chest radiography [8]. In addition, CT imaging can be used to identify lung atelectasis due to poor positioning of the endotracheal tube as well as potentially directing ventilation to achieve optimal pressures and air recruitment [8]. This is important in relation to COVID-19 since patients in ICU require optimisation of ventilatory settings and it is increasingly recognised that prone ventilation appears favourable [9]. Nevertheless, in the clinical setting, the benefits of routine CT imaging must be weighed against the considerable practicalities and risks, including that of infection transmission associated with transporting a patient from the intensive care unit, or elsewhere in the hospital, to the radiology department [8]. The Royal College of Radiologists does not recommend the use of CT-scanning as a diagnostic tool for COVID-19, as a negative CT result does not rule out COVID-19 infection, except for when CT-scanning results can change the management course, such as in those who require emergency surgery [10]. Furthermore, both the American Society of Emergency Radiology and the Society of Thoracic Radiology advise against the use of CT-scanning as a diagnostic tool for COVID-19 [11].

Given the recent observational studies of chest CT and RT-RCR in detecting COVID-19 cases [4,12-23], there is a need for ongoing systematic reviews and meta-analyses of studies that have directly compared the performance of these tests in the same patient cohorts. This study aimed to compare the performance of chest CT-scan versus RT-PCR for the diagnosis of COVID-19 by systematic review and meta-analysis of truly comparative studies.

## 2. Methods

A systematic review and meta-analysis were conducted as per the Preferred Reporting Items for Systematic Reviews and Meta-Analyses (PRISMA) guidelines [24].

### 2.1. Eligibility criteria

All relevant studies comparing chest CT with RT-PCR (as the reference standard) in the diagnosis of patients with suspected COVID-19 were included [4,12-23]. Chest CT was the intervention group of interest and RT-PCR was the comparator. All patients were included irrespective of age, gender or co-morbidity status. Studies that did not compare the two tests directly within the same patient cohort were excluded. Studies enrolling only confirmed COVID-19 cases or studies performing chest CT for prognostic purposes were also excluded.

### 2.2. Primary Outcomes

Sensitivity, specificity and accuracy for the diagnosis of COVID-19 cases were the primary outcomes. Sensitivity (true positive rate) is the proportion of actual COVID-19 cases a test correctly identifies. Specificity (true negative rate) is the proportion of non-COVID-19 cases a test correctly identifies. Accuracy is the proportion (true positives and true negatives) of total cases examined a test correctly identifies.

### 2.3. Secondary Outcomes

The secondary outcomes were positive predictive value (in the event of a positive test the probability the patient is truly positive), negative predictive value (in the event of a negative test the probability the patient is truly negative), positive (PLR) and negative likelihood (NLR) ratios (respectively, the probability a person who has COVID-19 testing positive divided by the probability of a person who does not have COVID-19 testing positive [PLR] and the probability of a person who has COVID-19 testing negative divided by the probability of a person who does not have COVID-19 testing negative [NLR]), and discrepancy of findings between both investigations.

### 2.4. Search Strategy

Three authors independently searched online databases, including MEDLINE, EMBASE, EMCARE, CINAHL and the Cochrane Central Register of Controlled Trials (CENTRAL). The last search was run on 23^rd^ of August 2020. Thesaurus headings, search operators and limits in each of the above databases were adapted accordingly. In addition, the WHO International Clinical Trials Registry, ClinicalTrials.gov, ISRCTN Register and medRxiv were also searched for ongoing and unpublished studies. The search terminologies included “COVID-19”, “Coronavirus”, “SARS-CoV-2”, “CT”, “computed tomography”, “PCR” and “polymerase chain reaction”. The bibliographic lists of relevant articles were also reviewed.

### 2.5. Selection of Studies

The title and abstract of articles identified from the literature searches were assessed. The full texts of relevant reports were retrieved and those articles that met the eligibility criteria of our review were selected.

### 2.6. Data Extraction

An electronic data extraction spreadsheet was created in line with Cochrane’s data collection form for intervention reviews. Three authors cooperatively collected and recorded the results with any disagreements resolved via discussion.

### 2.7. Data Synthesis

Data synthesis was conducted using the Review Manager 5.3 software and Microsoft Excel. The results are reported in forest plots with 95% Confidence Intervals (CIs) and tables. A summary receiver operator characteristic (sROC) curve was also generated.

### 2.8. Subgroup Analyses

Additional sub-group comparisons were performed to assess differences in results for China-based versus non-China and large versus small studies.

### 2.9. Methodological Quality and Risk of Bias Assessment

Three authors independently assessed the methodological quality and risk of bias of included studies using the Newcastle-Ottawa Scale [25]. It offers a maximum score of nine stars across three domains: selection, comparability and exposure. The overall rating of good, fair or poor quality was based on the Agency for Healthcare Research and Quality (AHRQ) standards [25].

## 3. Results

### 3.1. Literature Search Results

Our search strategy retrieved 1,630 studies. Thirteen studies met the eligibility criteria for inclusion following systematic review of abstracts and manuscripts as appropriate (Figure 1).

**Figure 1:**
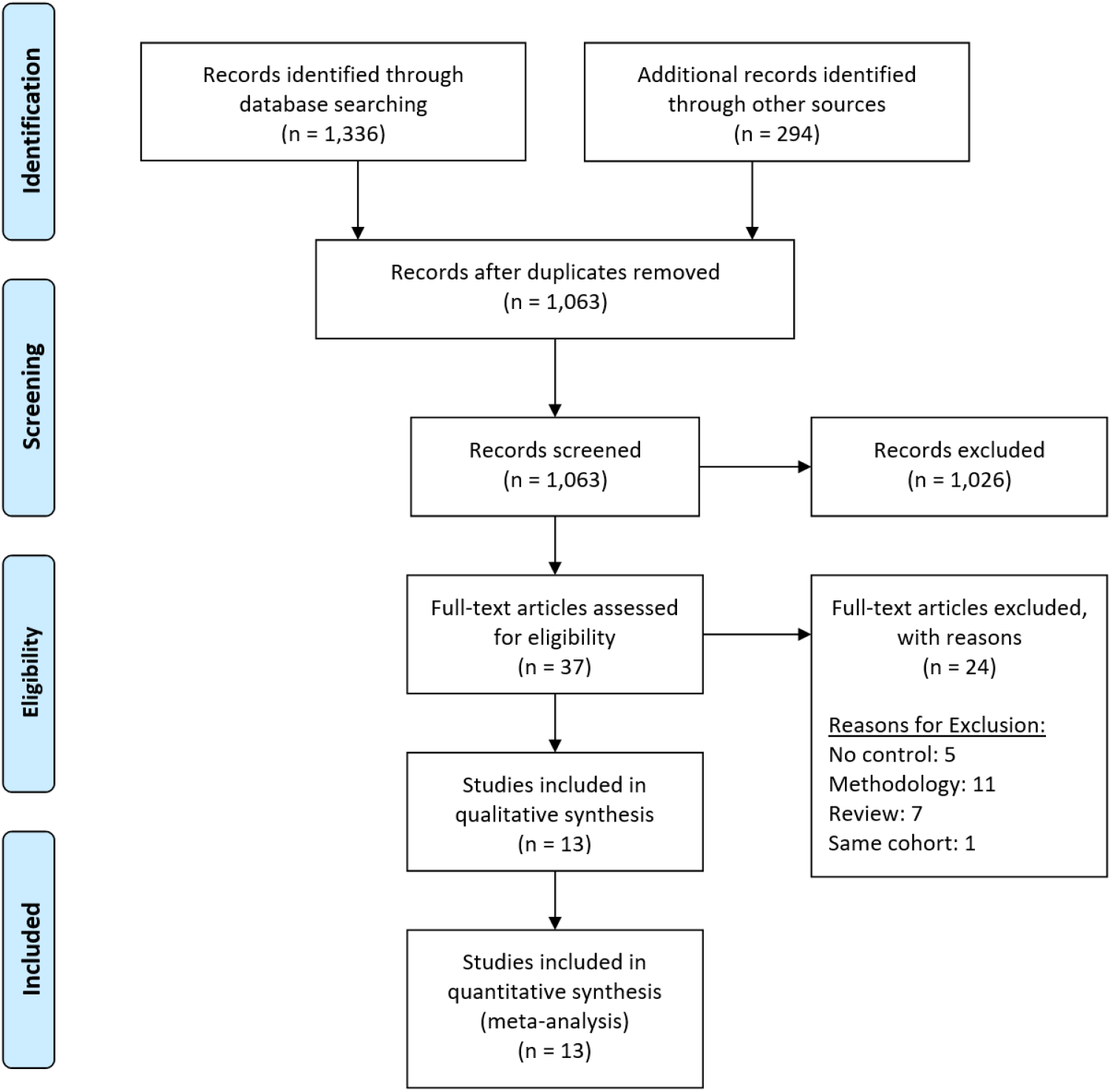
Prisma Flow Diagram. The PRISMA diagram details the search and selection processes applied during the overview. PRISMA, Preferred Reporting Items for Systematic Reviews and Meta-Analyses.

### 3.2. Characteristics of Included Studies

The baseline characteristics of the included studies are summarised in Table 1. All included studies were comparative with CT scans being assessed using RT-PCR as the reference (‘gold’) standard.

**Table 1.**
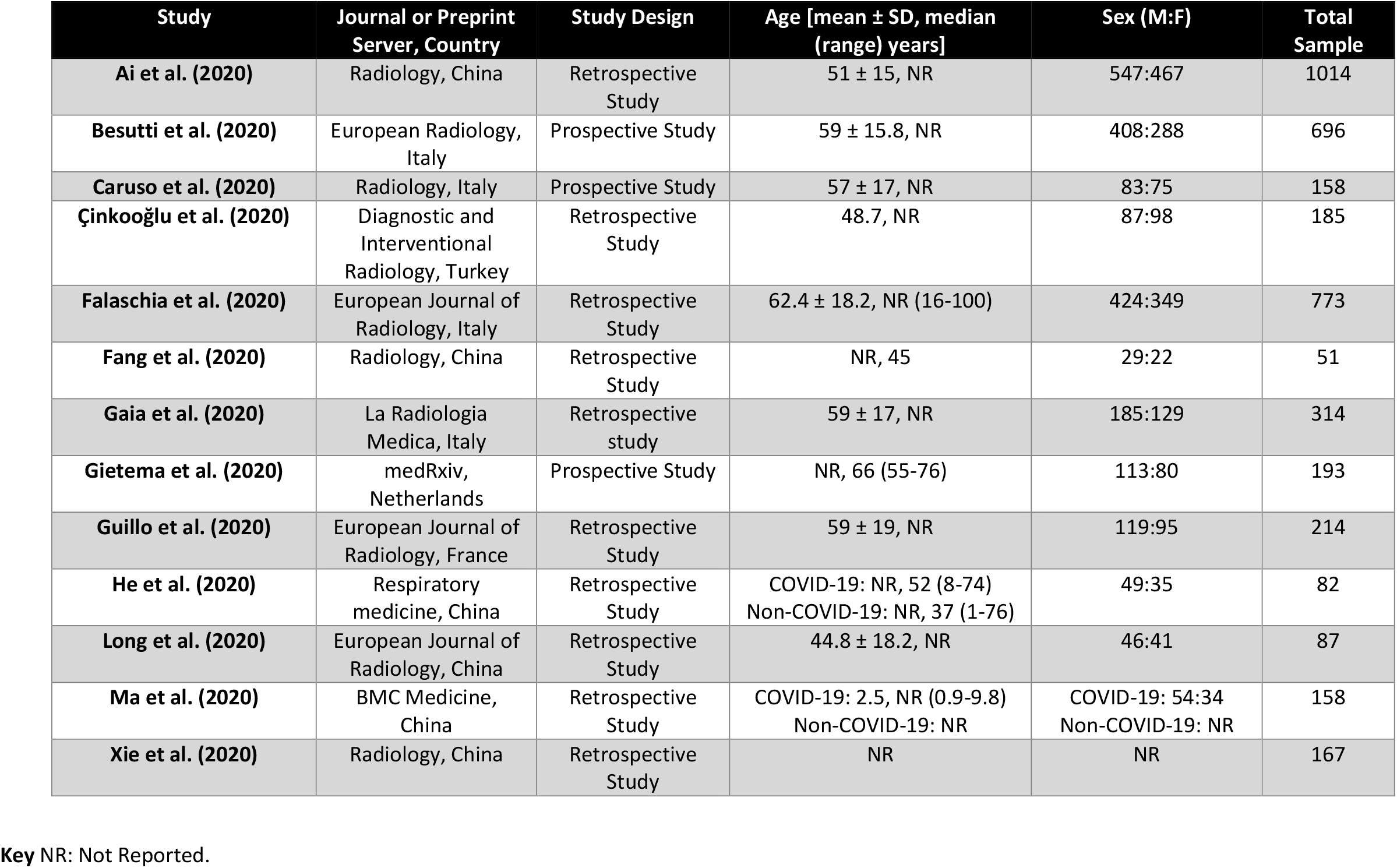
Baseline Characteristics of the Included Studies.

| Study | Journal or Preprint<br>Server, Country | Study Design | Age [mean ± SD, median<br>(range) years] | Sex (M:F) | Total<br>Sample |
| --- | --- | --- | --- | --- | --- |
| **Ai et al. (2020)** | Radiology, China | Retrospective<br>Study | 51 ± 15, NR | 547:467 | 1014 |
| **Besutti et al. (2020)** | European Radiology,<br>Italy | Prospective Study | 59 ± 15.8, NR | 408:288 | 696 |
| **Caruso et al. (2020)** | Radiology, Italy | Prospective Study | 57 ± 17, NR | 83:75 | 158 |
| **Çinkooğlu et al. (2020)** | Diagnostic and<br>Interventional<br>Radiology, Turkey | Retrospective<br>Study | 48.7, NR | 87:98 | 185 |
| **Falaschia et al. (2020)** | European Journal of<br>Radiology, Italy | Retrospective<br>Study | 62.4 ± 18.2, NR (16-100) | 424:349 | 773 |
| **Fang et al. (2020)** | Radiology, China | Retrospective<br>Study | NR, 45 | 29:22 | 51 |
| **Gaia et al. (2020)** | La Radiologia<br>Medica, Italy | Retrospective<br>study | 59 ± 17, NR | 185:129 | 314 |
| **Gietema et al. (2020)** | medRxiv,<br>Netherlands | Prospective Study | NR, 66 (55-76) | 113:80 | 193 |
| **Guillo et al. (2020)** | European Journal of<br>Radiology, France | Retrospective<br>Study | 59 ± 19, NR | 119:95 | 214 |
| **He et al. (2020)** | Respiratory<br>medicine, China | Retrospective<br>Study | COVID-19: NR, 52 (8-74)<br>Non-COVID-19: NR, 37 (1-76) | 49:35 | 82 |
| **Long et al. (2020)** | European Journal of<br>Radiology, China | Retrospective<br>Study | 44.8 ± 18.2, NR | 46:41 | 87 |
| **Ma et al. (2020)** | BMC Medicine,<br>China | Retrospective<br>Study | COVID-19: 2.5, NR (0.9-9.8)<br>Non-COVID-19: NR | COVID-19: 54:34<br>Non-COVID-19: NR | 158 |
| **Xie et al. (2020)** | Radiology, China | Retrospective<br>Study | NR | NR | 167 |

**Key** NR: Not Reported.

### 3.3. Primary Outcomes

#### 3.3.1. Discriminatory power (sensitivity and specificity)

Sensitivity to detect COVID-19 cases was reported in 13 studies enrolling 4092 patients (Figure 2). The median sensitivity of CT for the identification of COVID-19 cases was 0.91 (range 0.82-0.98) as demonstrated in Table 2. The specificity of CT scanning in identifying COVID-19 was reported in 10 studies enrolling 3689 patients (Figure 2). The median specificity for chest CT was 0.775 (range 0.25-1.00) as shown in Table 2. The discriminatory power of CT scanning in identifying patients with COVID-19 from non-infected patients, based on area under the sROC, is shown in Figure 3.

**Table 2.** The performance of CT scan for COVID-19 infection.

| Study ID | TP | FP | FN | TN | Sensitivity<br>[95% CI] | Specificity<br>[95% CI] | Accuracy | PPV | NPV | PLR | NLR |
| --- | --- | --- | --- | --- | --- | --- | --- | --- | --- | --- | --- |
| **Ai 2020** | 580 | 308 | 21 | 105 | 0.97<br>[0.95, 0.98] | 0.25<br>[0.21, 0.30] | 0.68 | 0.65 | 0.83 | 1.29 | 0.14 |
| **Besutti 2020** | 520 | 61 | 31 | 84 | 0.94<br>[0.92, 0.96] | 0.58<br>[0.49, 0.66] | 0.89 | 0.89 | 0.73 | 2.24 | 0.09 |
| **Caruso 2020** | 60 | 42 | 2 | 54 | 0.97<br>[0.89, 1.00] | 0.56<br>[0.46, 0.66] | 0.72 | 0.59 | 0.96 | 2.21 | 0.06 |
| **Çinkooğlu 2020** | 147 | 0* | 38 | 0 | 0.79<br>[0.73, 0.85] | Not estimable | 0.79 | 1.00 | 0 | Not estimable | Not estimable |
| **Falaschia 2020** | 419 | 66 | 43 | 245 | 0.91<br>[0.88, 0.93] | 0.79<br>[0.74, 0.83] | 0.86 | 0.86 | 0.85 | 4.27 | 0.12 |
| **Fang 2020** | 50 | 0* | 1 | 0 | 0.98<br>[0.90, 1.00] | Not estimable | 0.98 | 1.00 | 0 | Not estimable | Not estimable |
| **Gaia 2020** | 147 | 24 | 15 | 128 | 0.91<br>[0.85, 0.95] | 0.84<br>[0.77, 0.90] | 0.87 | 0.86 | 0.89 | 5.75 | 0.11 |
| **Gietema 2020** | 74 | 35 | 9 | 75 | 0.89<br>[0.80, 0.95] | 0.68<br>[0.59, 0.77] | 0.77 | 0.68 | 0.89 | 2.80 | 0.16 |
| **Guillo 2020** | 113 | 0 | 24 | 77 | 0.82<br>[0.75, 0.88] | 1.00<br>[0.95, 1.00] | 0.89 | 1.00 | 0.76 | Not estimable | 0.17 |
| **He 2020** | 26 | 2 | 8 | 46 | 0.76<br>[0.59, 0.89] | 0.96<br>[0.86, 0.99] | 0.88 | 0.93 | 0.85 | 18.35 | 0.25 |
| **Long 2020** | 35 | 0* | 1 | 51 | 0.97<br>[0.85, 1.00] | 1.00<br>[0.93, 1.00] | 0.99 | 1.00 | 0.98 | Not estimable | 0.03 |
| **Ma 2020** | 43 | 26 | 7 | 82 | 0.86<br>[0.73, 0.94] | 0.76<br>[0.67, 0.84] | 0.79 | 0.62 | 0.92 | 3.57 | 0.18 |
| **Xie 2020** | 160 | 0* | 7 | 0 | 0.96<br>[0.92, 0.98] | Not estimable | 0.96 | 1.00 | 0 | Not estimable | Not estimable |
| Median (Range) |  |  |  |  | 0.91<br>(0.82-0.98) | 0.775<br>(0.25-1.00) | 0.87<br>(0.68-0.99) | 0.89<br>(0.59-1.00) | 0.85<br>(0-0.98) | 3.185<br>(1.29-18.35) | 0.13<br>(0.03-0.25) |

**Key** TP: True Positive, FP: False Positive, FN: False Negative, TN: True Negative, CI: Confidence Interval, PPV: Positive Predictive Value, NPV: Negative Predictive Value, PLR: Positive Likelihood Ratio, NLR: Negative Likelihood Ratio. \* Assumes no false positive due to non-reported data.

**Figure 2:**
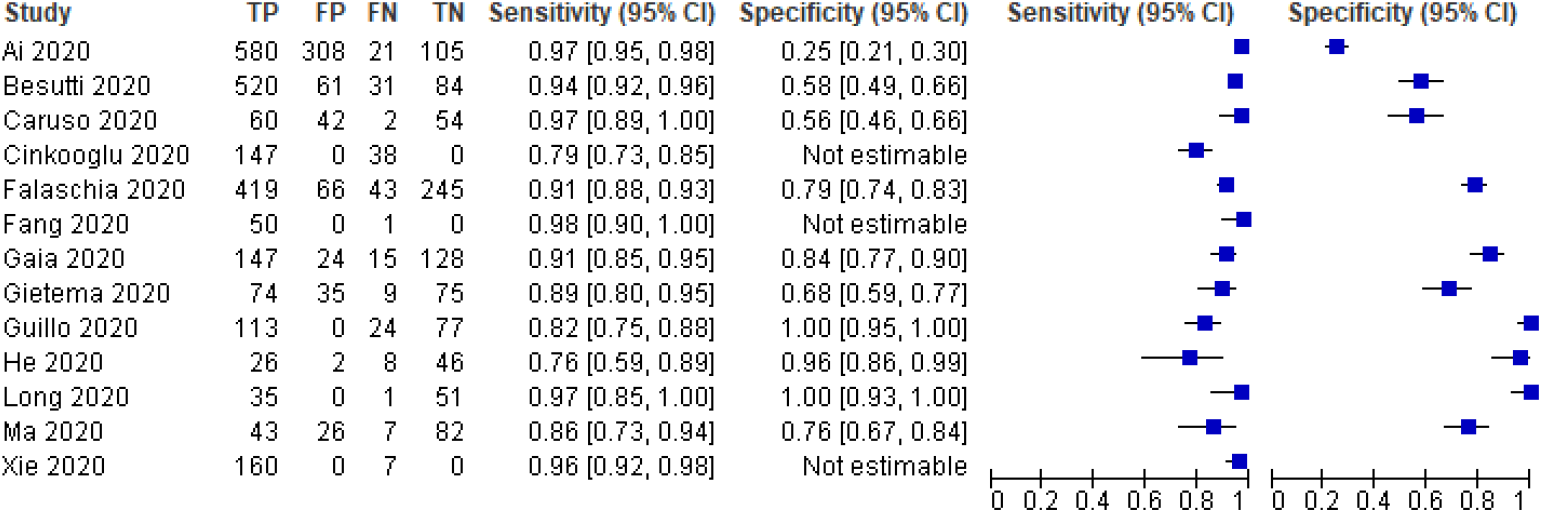
Forest Plot of Chest CT – Sensitivity and Specificity for the identification of COVID-19 cases. **Key** CT: Computed Tomography, COVID-19: Coronavirus Disease 2019.

**Figure 3:**
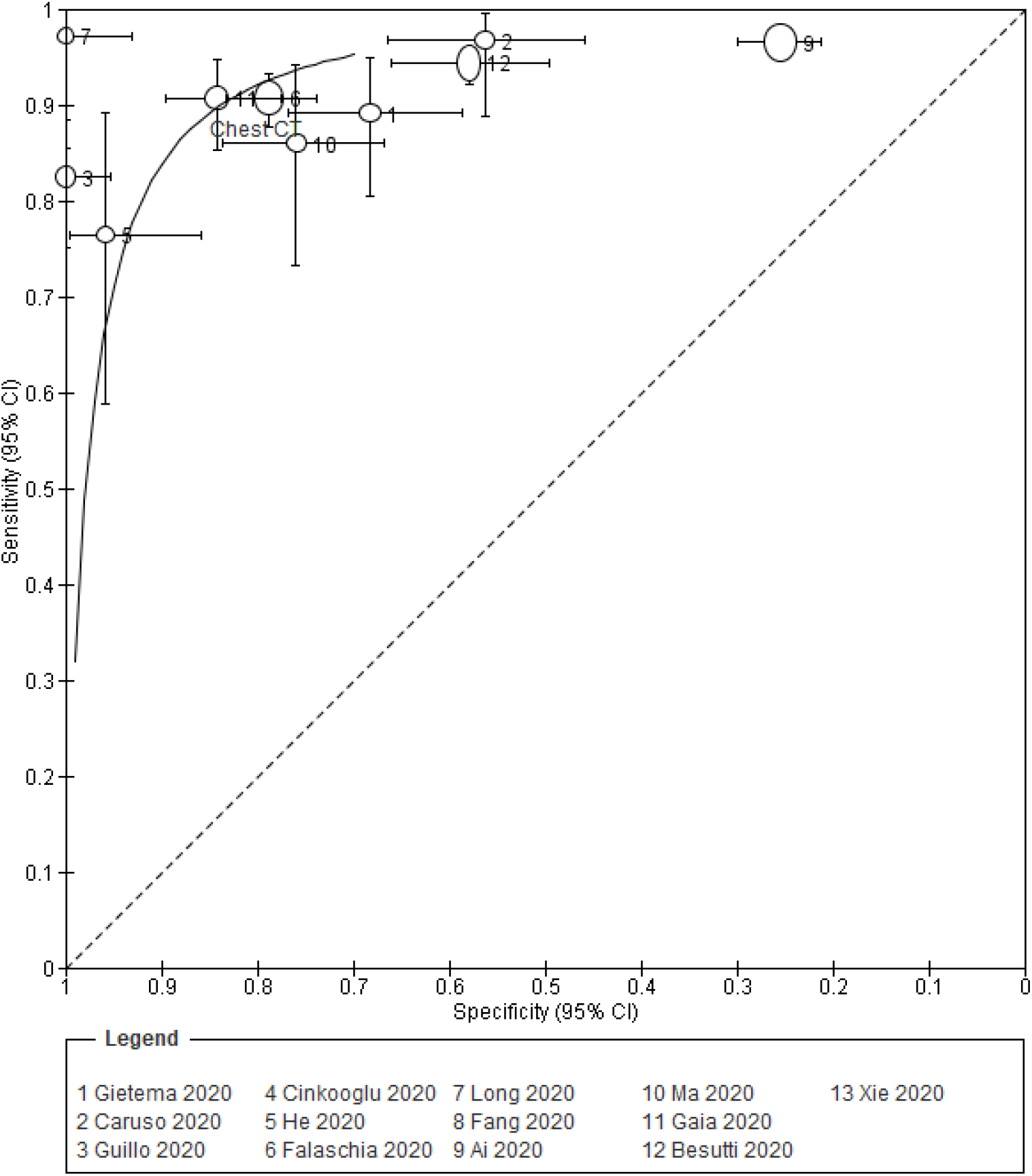
Summary Receiver Operator Characteristic (sROC) curves for the performance of Chest CT in the initial diagnosis of COVID-19. **Key** CT: Computed Tomography

#### 3.3.2. Diagnostic effectiveness (accuracy)

Accuracy to detect and exclude COVID-19 cases by CT scan was reported in 13 studies enrolling 4092 patients with a median accuracy of 0.87 (range 0.68-0.99) as demonstrated in Table 2.

### 3.4. Secondary Outcomes

#### 3.4.1. Other test performance characteristics

As shown in Table 2, CT scanning had median positive and negative predictive values of 0.89 (range 0.59-1.00) and 0.85 (range 0.73-0.98), respectively. Additionally, the median PLR and NLR for CT scan were 3.185 (range 1.29-18.35) and 0.13 (range 0.03-0.25) in turn.

#### 3.4.2. Discrepancy of Findings

Ai et al. [4] (n = 1014) reported that 75% (n = 310) of patients with negative RT-PCR results and 97% (n = 583) of those with positive results had positive chest CT findings. Across all studies, 9.1% (230/2529) of PCR positive patients had a negative CT scan and 79.2% (42/53) of patients who initially had a negative PCR but then had a positive PCR later had a CT scan suggestive of COVID-19 (Supplemental Material 1).

### 3.5. Subgroup Analyses

#### 3.5.1. China and Non-China Based Studies

China-based studies had an accuracy range of 0.68 to 0.99 (median 0.92; 50% of studies had an accuracy of >0.95) versus 0.72 to 0.89 (median 0.86; no studies had an accuracy of >0.9) in non-China studies in the identification of COVID-19 (Supplemental Material 2).

#### 3.5.2. Larger and Smaller Studies

There was also a difference between larger (n ≥100) and smaller (n <100) study subgroups in accuracy for the identification COVID-19 (Supplemental Material 3). Larger studies had an accuracy range of 0.68 to 0.96 (median 0.825; 30% of studies had an accuracy ≥0.88) versus 0.88 to 0.99 (median 0.98; all studies had an accuracy of ≥0.88) for smaller studies.

### 3.6. Methodological Quality and Risk of Bias Assessment

Although comparability was low in all the studies included, selection and exposure were of higher quality (Supplemental Material 4). Overall, all studies were of good quality based on the AHRQ standards [25].

## 4. Discussion

Our findings suggest that CT scanning performs relatively well compared to RT-PCR as the reference standard in the initial detection of COVID-19 in the clinical context of the studies presented. Relatively few false negatives (approximately 9%) can be expected with CT scanning in the context of a pandemic, although the expected false positive rate is higher at approximately 22.5%. The performance characteristics of all tests is context dependent, however, which means that the performance of CT for identifying COVID-19 will be different outside of a pandemic and in areas of the world with different endemicities. When the causes of bilateral CT changes return to the usual heterogeneous state, with SARS-CoV-2 only one of many potential causes, as was the situation pre-pandemic, the performance of CT will likely deteriorate with an even higher number of false positives, in keeping with the relatively modest PLR for CT, even within a pandemic setting [4]. For patients requiring hospitalisation with an acute cardio-respiratory presentation who undergo CT scanning, however, a negative CT scan is reassuring, particularly if RT-PCR is also negative in keeping with the low NLR for CT. Our results cannot be extrapolated to healthier SARS-CoV-2 patient cohorts who are managed on an outpatient basis or in a community setting.

Diagnostic tests always have inherent strengths and weaknesses. In the case of the COVID-19 pandemic it would be preferable to have a simple, rapid test with high sensitivity leading to a low number of false negatives thereby allowing accurate decision-making regarding, for example, who may be infectious and requires isolation in the hospital setting or home isolation in the community. Based on those who had a suggestive CT but tested negative with RT-PCR, our findings suggest that CT scan may be able to detect a high proportion of hospitalised COVID-19 cases overall and may be useful when PCR testing is initially negative but clinical suspicion high [4]. There were relatively few patients who had a negative CT scan, but who were initially RT-PCR positive. Of the studies that reported patients who were initially RT-PCR negative, but subsequently became positive, 79.2% of patients (42/53) had a suggestive CT. It is important to note that across the studies, however, only a small proportion who were initially RT-PCR negative were systematically retested.

Overall, our results suggest that chest CT scan may be a useful adjunct to RT-PCR in the initial detection of COVID-19 in certain circumstances, but not routinely, which supports published guidance and thus we do not advocate a change in practice. CT scans are subject to their inherent limitations. Firstly, they are best avoided during pregnancy due to the excessive radiation and risk of harming the foetus [26]. In addition, over-scanning patients with suspected COVID-19 can increase the risk of incidental imaging findings and an unnecessary financial burden on the healthcare system [27]. Furthermore, the use of CT-scans for suspected cases is not practical since SARS-CoV-2 has a relatively high transmission rate; each CT machine needs to be sterilised after use, which can waste time and resources as well as overworking the radiology department in times of need resulting in delayed reporting of other potentially life-threatening conditions.

Given the above, and based on our results therefore, we suggest that chest CT is limited to a supplementary diagnostic role for COVID-19 in patients with suspicious symptoms, but who have had at least one negative RT-PCR test result. In such situations, a repeat upper respiratory tract RT-PCR, which is simple to do, rather than a CT scan should be performed first, although in our clinical experience some patients with suspicious presentations are repeatedly negative or only positive on a third or fourth test. Lower respiratory tract specimens (i.e. sputum or bronchoalveolar lavage) are more likely to be positive in this patient cohort, although most hospitalised patients are neither productive nor ventilated.

This review provides a summary of the best available evidence using a systematic approach and evaluated the risk of bias of included studies [4,12-23]. Although Kim et al. recently published a meta-analysis on this topic, many of the studies included were not comparative (i.e. the tests were not compared within the same patient cohorts) [28]. Our study synthesised comparative studies only, and also included additional recently published studies and those in preprint, so our results may more accurately represent how the investigations can be expected to perform and compare to RT-PCR in routine clinical practice and as the pandemic progresses. Additionally, we also performed important sub-group analyses that demonstrate how these tests may compare in different contexts, including the fallacy of basing conclusions about a test’s performance early in a pandemic on small studies in one geographic location.

The reported results should be considered in the context of inherent limitations. Only 13 non-randomised studies were identified enrolling a total of 4092 participants, which may not be enough to draw definite conclusions and suggesting the need for further robust prospective diagnostic research in COVID-19. The performance of molecular tests for SARS-CoV-2 is likely to improve and be closer at the point-of-care with time. Likewise, the use of, for example, machine learning algorithms to interpret chest CT scan appearances may more accurately confirm COVID-19 than human reporting in the future, and thereby decrease the false-positive rate of CT scanning that can be expected to increase as the pandemic wanes. Some included studies had incomplete data resulting in difficulty in comparing both interventions completely. Additionally, all but one study included a relatively small number of patients and the majority were of retrospective observational design, with all the associated caveats. It is likely that further large comparative, and possibly prospective, studies will be reported with further meta-analyses required. The between-study heterogeneity for outcomes was high for sensitivity and accuracy.

## 5. Conclusions

Our study synthesised the best available comparative evidence, using a systematic approach, for the identification of COVID-19 using chest CT versus RT-PCR as the reference standard [4,12-23]. Our results suggest that chest CT scan may be a useful adjunct in the initial detection of COVID-19 in certain hospitalised patients in the context of a pandemic. We suggest RT-PCR remains the primary diagnostic tool and that chest CT is only considered if there is a strong clinical suspicion of COVID-19 with repeatedly negative RT-PCR test results, providing infection prevention and control measures can be maintained. A negative CT scan is likely to be reassuring in a RT-PCR negative patient requiring hospitalisation for possible COVID-19.

## Data Availability

The author(s) declare(s) that they had full access to all of the data in this study and the author(s) take(s) complete responsibility for the integrity of the data and the accuracy of the data analysis.

## 6. Declarations

### Ethics Approval and Consent to Participate

Not Applicable.

### Consent for Publication

Not Applicable.

### Availability of Data and Materials

The datasets generated and analysed during the current study are available from the corresponding author on reasonable request.

### Authors’ Contributions

MK contributed in study concept and design as well as acquisition, analysis and interpretation of data. A Abul, AH, A Alsaif helped in collecting and analysing data as well as drafting the manuscript. SA and MA were responsible for screening additional papers and drafting the manuscript. SA, MA and A Alsaif assessed the quality of included studies. GB was the lead supervisor for critically reviewing the project, including editing of manuscript. All authors read and approved the final manuscript.

## Acknowledgements

Not Applicable.

## Supplemental Material 1

**Table 1.** Additional data. Column 1 shows the percentage of PCR positive patients who have a negative CT scan. Column 2 shows the proportion of patients with a CT scan suggestive of COVID-19 in patients who initially had a negative PCR, but then later had a positive PCR.

| Study ID | Number of Patients with Positive PCR but Negative CT Scan ÷ Total Number of PCR Positive Patients (%) | Number of patients with CT changes from population with initial negative PCR that then became PCR positive later ÷ Number of patients with initial negative but then positive PCR later (%) |
| --- | --- | --- |
| Ai 2020 | 21/601 (3.5%) | 10/15 (67%) |
| Besutti 2020 | 31/551 (6.1%) | 5/11 (45.4%) |
| Caruso 2020 | 2/62 (3.2%) | NR |
| Çinkooğlu 2020 | 38/174 (21.8%) | 10/10 (100%) |
| Falaschia 2020 | 66/462 (14.3%) | NR |
| Fang 2020 | 1/36 (2.8%) | NR |
| Gaia 2020 | 15/162 (9.3%) | 6/6 (100%) |
| Gietema 2020 | 9/83 (10.8%) | NR |
| Guillo 2020 | 24/129 (18.6%) | NR |
| He 2020 | 8/27 (29.6%) | NR |
| Long 2020 | 1/30 (3.3%) | 6/6 (100%) |
| Ma 2020 | 7/50 (14%) | NR |
| Xie 2020 | 7/162 (4.3%) | 5/5 (100%) |
| Totals | 230/2529 (9.1%) | 42/53 (79.2%) |

**Key** NR: Not Reported

## Supplemental Material 2

**Table 2.**
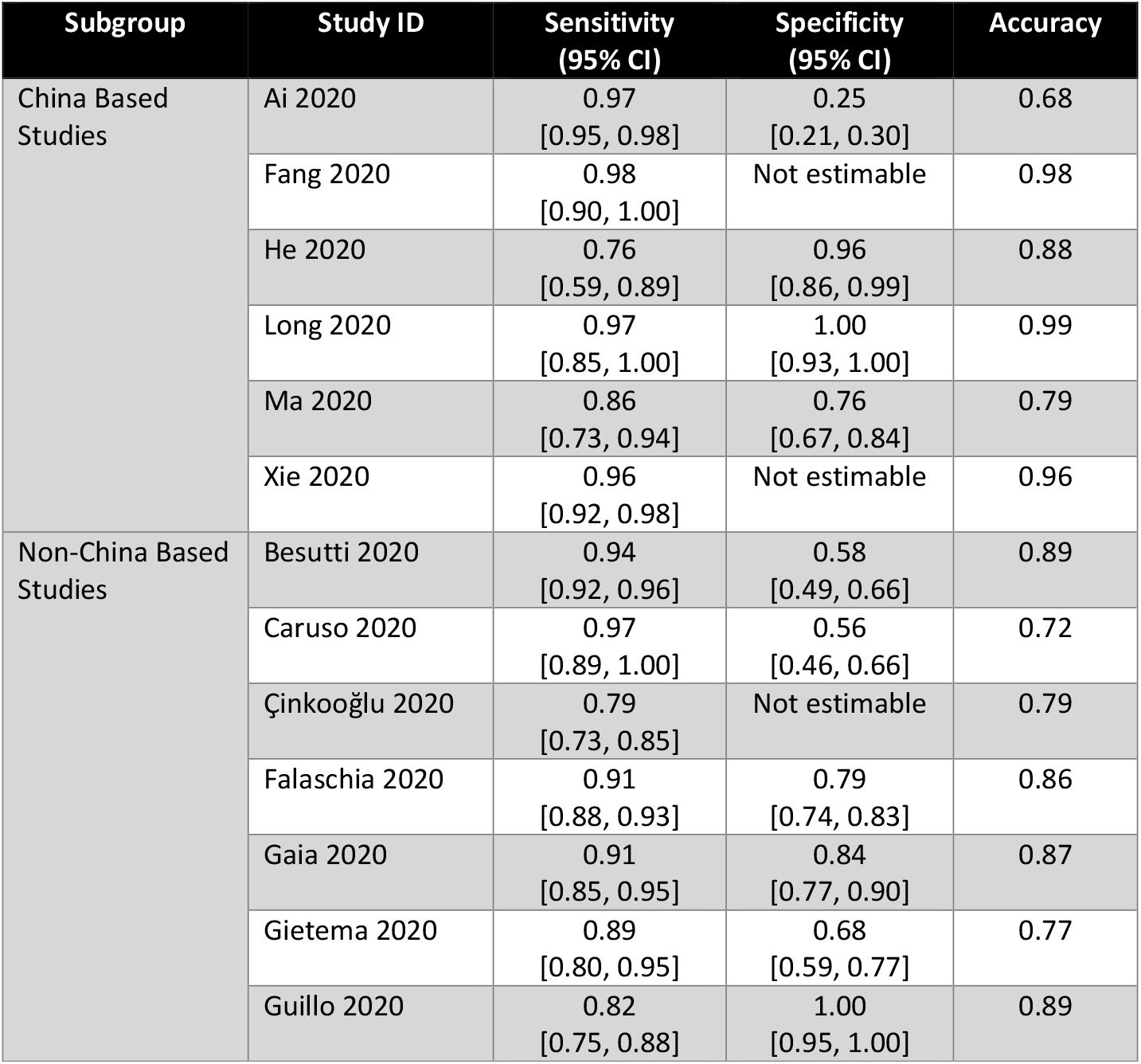
Subgroup Analysis of China vs. Non-China Based Studies for the performance of CT scan for COVID-19 infection.

## Supplemental Material 3

**Table 3.**
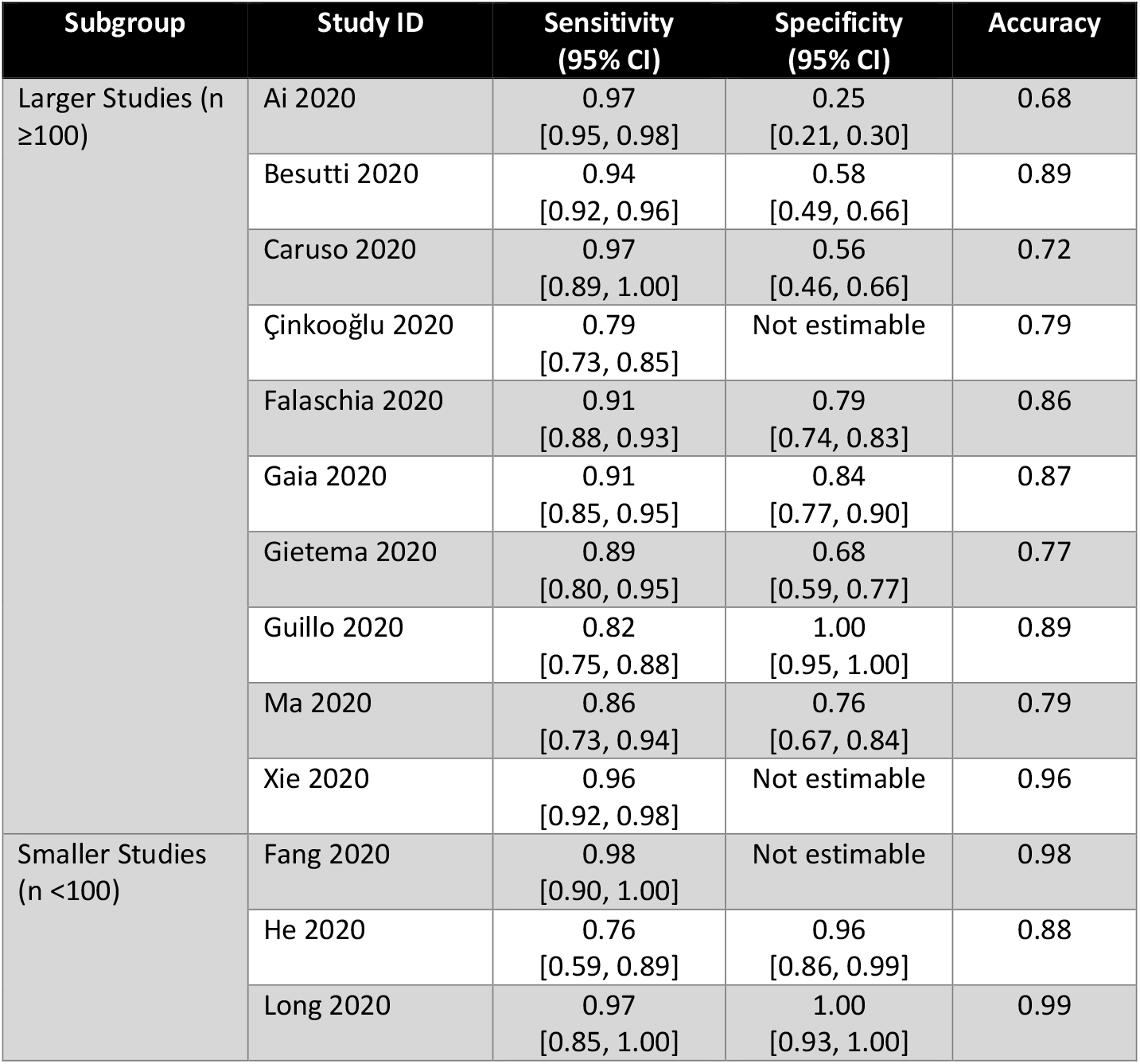
Subgroup Analysis of Larger vs. Smaller Studies for the performance of CT scan for COVID-19 infection.

## Supplemental Material 4

**Table 4.** Newcastle-Ottawa Scale to Assess the Quality of Non-Randomised Studies.

| **Study**<br>**(Year)** | **Selection**<br>**(out of 4)** | **Comparability**<br>**(out of 2)** | **Exposure**<br>**(out of 3)** |
| --- | --- | --- | --- |
| **Ai et al. (2020)** | **** | * | ** |
| **Besutti et al. (2020)** | *** | * | ** |
| **Caruso et al. (2020)** | **** | * | ** |
| **Çinkooğlu et al. (2020)** | *** | * | ** |
| **Falaschia et al. (2020)** | **** | * | ** |
| **Fang et al. (2020)** | *** | * | ** |
| **Gaia et al. (2020)** | *** | * | ** |
| **Gietema et al. (2020)** | **** | * | *** |
| **Guillo et al. (2020)** | **** | * | *** |
| **He et al. (2020)** | *** | * | ** |
| **Long et al. (2020)** | **** | * | ** |
| **Ma et al. (2020)** | **** | * | *** |
| **Xie et al. (2020)** | *** | * | *** |

